# Frontline healthcare workers’ experiences with personal protective equipment during the COVID-19 pandemic in the UK: a rapid qualitative appraisal

**DOI:** 10.1101/2020.10.12.20211482

**Authors:** Katarina Hoernke, Nehla Djellouli, Lily Jay Andrews, Sasha Lewis-Jackson, Louisa Manby, Sam Martin, Samantha Vanderslott, Cecilia Vindrola-Padros

**Author notes:** Correspondence to: K Hoernke. Copyright The Corresponding Author (KH) has the right to grant on behalf of all authors and does grant on behalf of all authors, an exclusive licence (or non exclusive for government employees) on a worldwide basis to the BMJ Publishing Group Ltd to permit this article (if accepted) to be published in BMJ editions and any other BMJPGL products and sublicences such use and exploit all subsidiary rights, as set out in our licence.

## Abstract

**Objectives:** To report frontline healthcare workers’ (HCWs) experiences with personal protective equipment (PPE) during the COVID-19 pandemic in the UK. To understand HCWs’ fears and concerns surrounding PPE, their experiences following its guidance and how these affected their perceived ability to deliver care during the COVID-19 pandemic.

**Methods:** A rapid qualitative appraisal study combining three sources of data: semi-structured in-depth telephone interviews with frontline HCWs (n=46), media reports (n=39 newspaper articles and 145,000 social media posts) and government PPE policies (n=25). HCWs interviewed were from secondary care, primary care and specialist community clinics. Media and policy data were from across the UK.

**Results:** A major concern was running out of PPE, putting HCWs and patients at risk of infection. Following national-level guidance was often not feasible when there were shortages, leading to re-use and improvisation of PPE. Frequently changing guidelines generated confusion and distrust. PPE was reserved for high-risk secondary care settings and this translated into HCWs outside these settings feeling inadequately protected. Participants were concerned about inequitable access to PPE for community, lower seniority, female and ethnic minority HCWs. Participants continued delivering care despite the physical discomfort, practical problems and communication barriers associated with PPE use.

**Conclusion:** This study found that frontline HCWs persisted in caring for their patients despite multiple challenges including inappropriate provision of PPE, inadequate training and inconsistent guidance. In order to effectively care for patients during the COVID-19 pandemic, frontline HCWs need appropriate provision of PPE, training in its use, as well as comprehensive and consistent guidance. These needs must be addressed in order to protect the health and well-being of the most valuable healthcare resource in the COVID-19 pandemic: our HCWs.

What is already known?

– PPE is an important component of infection prevention and control to protect HCWs delivering care on the frontline of an infectious disease outbreak.
– Frontline HCWs have reported challenges delivering care in PPE during the COVID-19 pandemic.
– Research understanding how HCWs responded to these challenges are lacking.

What are the new findings?

– HCWs faced multiple challenges delivering care including inadequate provision of PPE, inconsistent guidance and lack of training in its use.
– HCWs persisted delivering care despite the negative physical effects, practical problems, lack of protected time for breaks and communication barriers associated with wearing PPE.
– In the face of training, guidance and procurement gaps, HCWs improvised by developing their own informal communication channels to share information, they trained each other and bought their own PPE.
– HCWs reported inequalities accessing PPE based on the healthcare sector, gender, level of seniority and ethnicity.

What do the new findings imply?

– To feel safe and confident caring for patients, frontline HCWs need to be provided with appropriate size, quality and level of PPE, as well as training in its use.
– PPE guidance should be consistent, clearly communicated, and reflect the most up-to-date evidence-base for the safest level of PPE.
– Regular breaks for staff working in full PPE should be prioritised even in contexts of understaffing and PPE shortages as these are key aspects of well-being.

## INTRODUCTION

The provision of personal protective equipment (PPE) for frontline healthcare workers (HCWs) has become a defining problem of the Coronavirus disease 2019 (COVID-19) pandemic.^1^ The demand for PPE has put global supply chains under unprecedented strain.^2^ By March 2020, the World Health Organisation (WHO) called for rational PPE use and for global PPE manufacturing to be scaled up by 40%.^3^ This has led to widespread concerns regarding inadequate provision of PPE and its impact on the protection of frontline HCWs. In an international survey in April 2020, over half of HCWs had experienced PPE shortages, nearly a third were reusing PPE and less than half had adequate fit-testing.^4^ In the United Kingdom (UK), a third of respondents from a Royal College of Nurses (RCN) survey^5^ and over half from a British Medical Association (BMA) survey^6^ said they felt pressure to work without adequate PPE. Both surveys also raised concerns that Black, Asian and Minority Ethnic (BAME) and female HCWs may be disproportionately affected by PPE shortages. Additional concerns over impaired communication, physical discomfort, overheating and dehydration associated with PPE have also been raised.^7^ As of 20 July 2020, 313 HCWs had died from COVID-19 in the UK.^8^

Research on the appropriate level of PPE for COVID-19 is still ongoing.^9^ Severe Acute Respiratory Syndrome Coronavirus-2 (SARS-CoV-2) is thought to be transmitted via respiratory, contact and airborne transmission.^10^ Respiratory and contact precautions recommended by Public Health England (PHE) when caring for suspected cases include a Fluid-Resistant (Type IIR) Surgical Face Mask (FRSM), apron, gloves, and eye protection upon risk assessment.^11^ Airborne precautions recommended when caring for patients requiring aerosol generating procedures (AGPs) are higher and include a filtering facepiece 3 (FFP3) respirator, long-sleeved disposable fluid-repellent gown, gloves and eye protection.^11^

Knowledge from previous epidemics highlights the importance of PPE for frontline HCWs to reduce the spread of disease, safeguard HCWs’ health and well-being, and maintain a sustainable health workforce to curb the outbreak.^12^ Adequate provision of PPE, as well as clear guidance and training in its use help HCWs feel confident and prepared to deliver care.^13^ Previous epidemic research also highlights the value of understanding HCWs’ fears and concerns in order to support them on the frontline of an outbreak.^14^ Qualitative research methodologies are increasingly being used to inform response efforts. In the 2014 Ebola and 2015-16 Zika outbreaks, qualitative research helped generate context-specific, real-time recommendations to improve the planning and implementation of response efforts.^15^

PPE has become a critical issue for frontline HCWs in the COVID-19 pandemic but studies capturing HCWs’ experiences with PPE are lacking.^16^ The aims of this study were to determine (a) frontline HCWs’ experiences following local level (e.g. Trust) and national level (e.g. government) PPE guidance; (b) concerns and fears among HCWs regarding PPE in the context of the COVID-19 pandemic; and (c) how these experiences and concerns affected HCWs’ perceived ability to deliver care during the pandemic.

## METHODS

### Design

This study was part of a larger ongoing study on frontline HCWs’ perceptions and experiences of care delivery during the UK COVID-19 pandemic.^17^ We utilised a rapid appraisal methodology with three sources of data, including telephone interviews with frontline staff, a policy review, and media analysis (see Table 1). A rapid qualitative appraisal is an iterative approach to data collection and analysis, which triangulates findings between multiple sources of data to develop an understanding of a situation.^18^ It was chosen for its ability to generate targeted research in a timely manner in order to help inform response efforts to complex health emergencies.^15^ The use of an intensive, team-based approach with multiple sources of data helped to increase insight and validity of results.^19^

**Table 1:**
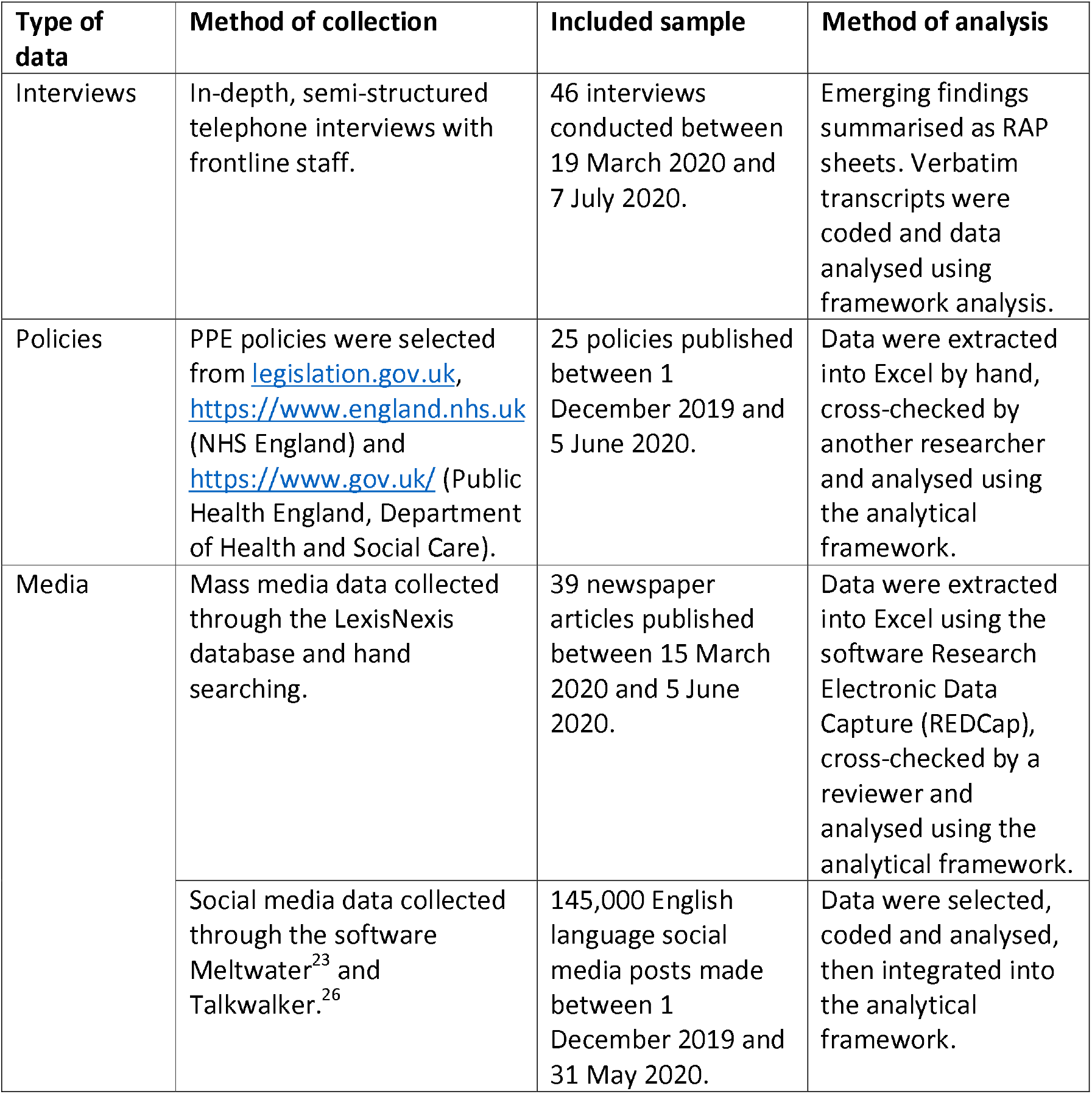
Methods of data collection and analysis.

### Sampling and recruitment

Participants were purposively sampled from Intensive Care Units (ICUs), Intensive Therapy Units (ITU), emergency departments, primary care and community clinics. They had a variety of experience, ranging from newly qualified to over 40 years working in the National Health Service (NHS). Participants were approached by clinical leads in their Trusts to gather verbal consent for the research team to contact them via email. Participants were provided with a participant information sheet and after filling out a consent form, had a telephone interview arranged.

### Data collection

Table 1 details data collection methods.

#### Interviews

46 in-depth, semi-structured telephone interviews with frontline HCWs were carried out using a broad topic guide. A multidisciplinary research team (including CVP, KH, LM and SLJ) conducted the interviews. Informed, written consent was obtained from all participants. Interviews were audio recorded, transcribed verbatim and all data anonymised. Emerging findings were summarised in the form of Rapid Assessment Process (RAP) sheets^18^ to increase familiarisation and engagement with the data.^20^ Interviews were included until data reached saturation, determined by no new themes emerging from RAP sheets.^21^

#### Policies

A review of 25 UK government policies and guidelines relating to PPE was carried out to contextualise HCWs’ experiences following PPE guidance using Tricco et al.’s framework.^22^ SLJ, LM and KH selected policies that met the inclusion criteria (see Appendix 1), cross-checked and extracted data into Excel.

#### Media

A rapid evidence synthesis of 39 newspaper articles and 145,000 English language Twitter posts meeting the inclusion criteria (see Appendix 1) was carried out utilising the same methodology as the policy review.^22^ LJA screened titles and full texts of mass media data with exclusions cross-checked by another researcher. SM and SV utilised the media monitoring software Meltwater^23^ to collect social media data using keyword searches on Twitter.

### Data analysis

The study was informed by a theoretical framework derived from anthropological perspectives on the material politics of epidemic responses.^24^ All streams of data were analysed using the Framework Method,^25^ as this type of analysis has been effective for rapid qualitative appraisals in previous epidemics.^15^ Social media data underwent additional demographic, discourse and sentiment analysis using the software TalkWalker.^26^ All sources of data were coded with the same analytical framework to triangulate findings between the different streams of data.

## RESULTS

### Participants

Participants represented a range of HCWs. The majority were doctors and nurses working in hospital settings and one non-HCW was included for their expertise in IPC services. (Table 2).

**Table 2:** Participant demographics.

|  | Sample (n=46) | Percentage total (%) |
| --- | --- | --- |
| **Role** |  |  |
| Doctor | 28 | 60.87% |
| Nurse | 8 | 17.39% |
| Healthcare practitioner | 3 | 6.52% |
| Pharmacist | 2 | 4.35% |
| Physician associate | 1 | 2.17% |
| Dietician | 1 | 2.17% |
| Speech and language therapist | 1 | 2.17% |
| Healthcare assistant | 1 | 2.17% |
| Infection Prevention and Control Service | 1 | 2.17% |
| **Sector** |  |  |
| Secondary care (hospital) | 40 | 87.0% |
| Primary care (general practice) | 4 | 8.7% |
| Specialist community services | 2 | 4.3% |
| **Ethnicity** |  |  |
| White | 40 | 86.96% |
| BAME | 6 | 13.04% |
| **Total** | **46** | **100%** |

**Table 3:**
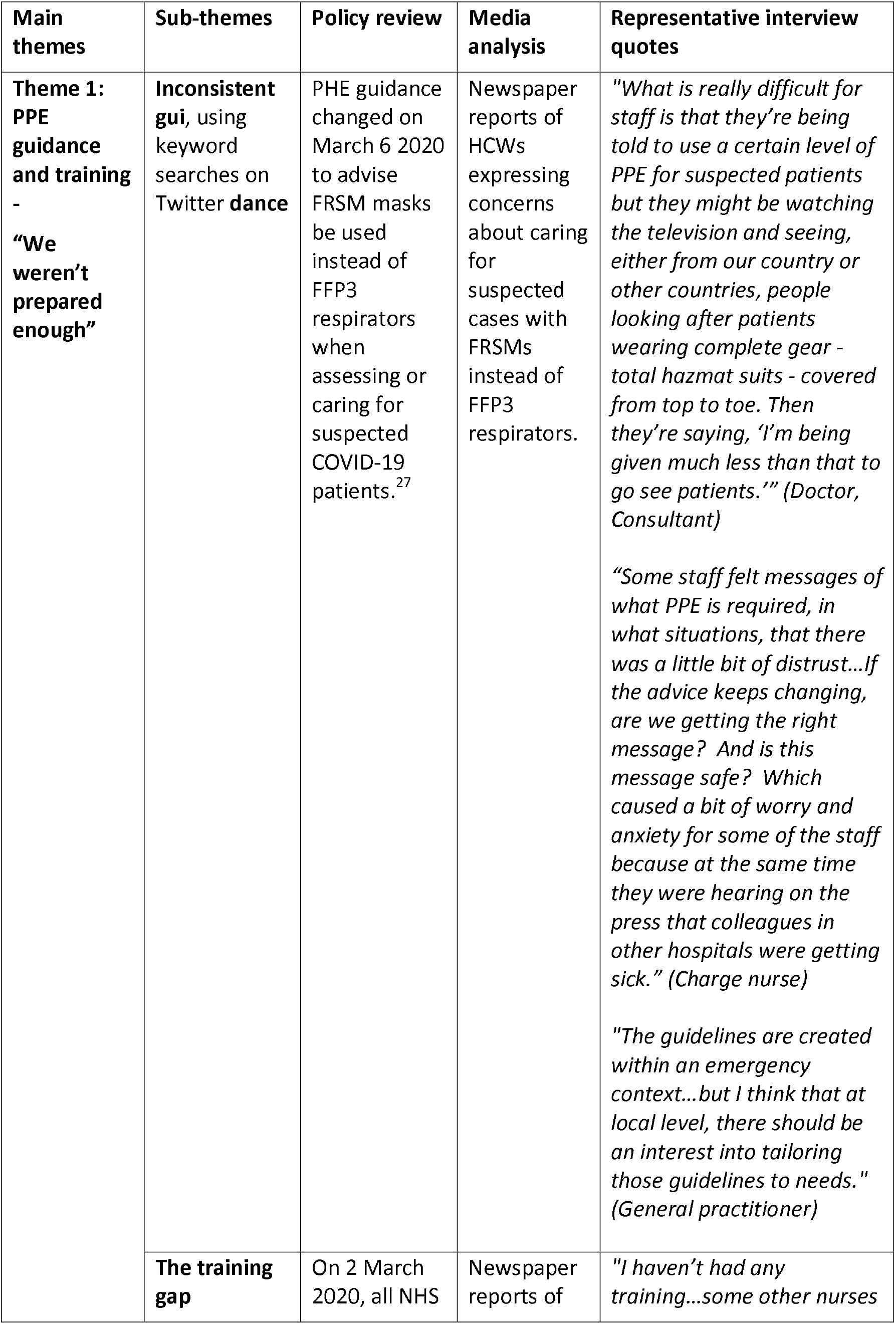
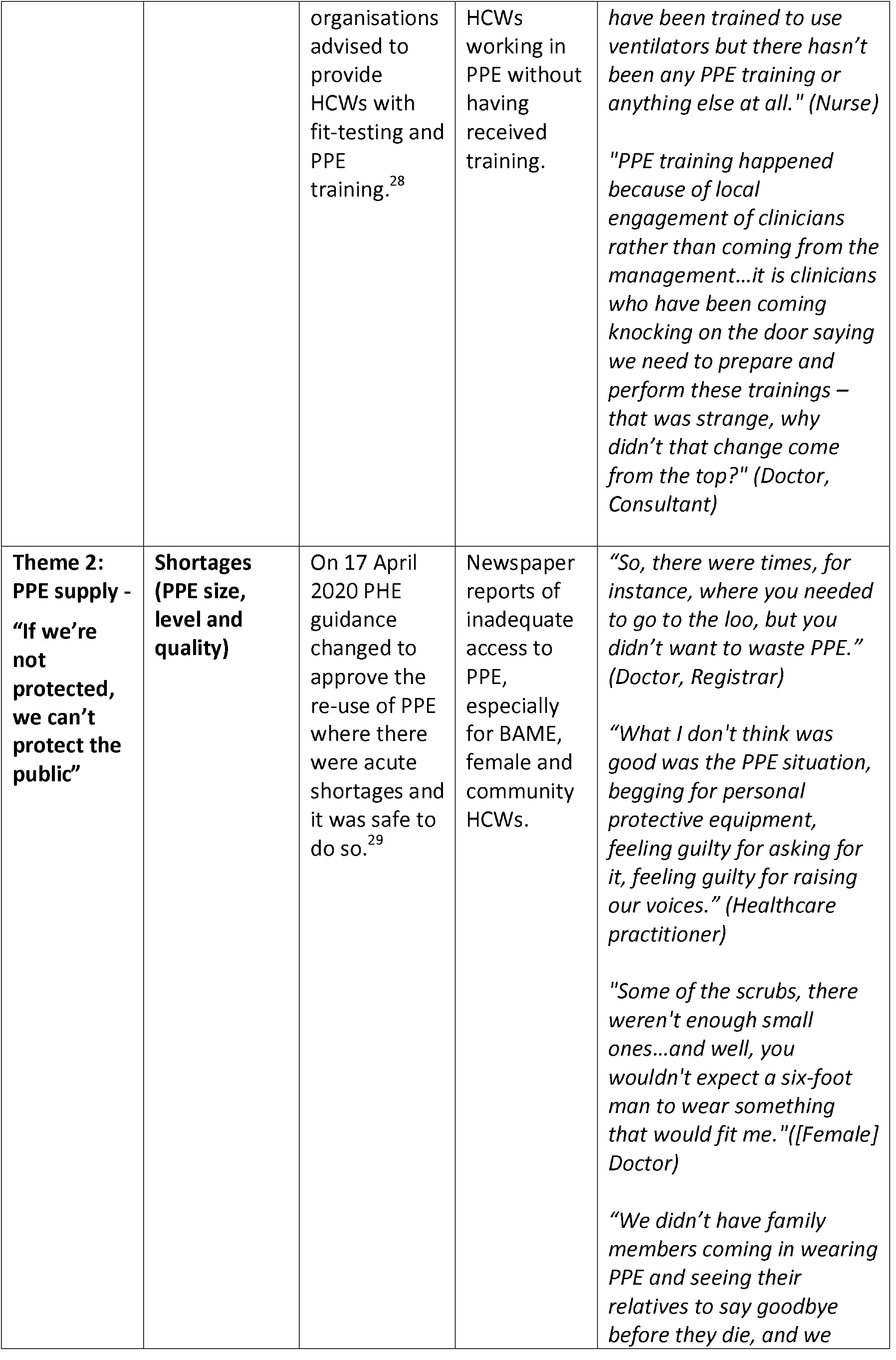
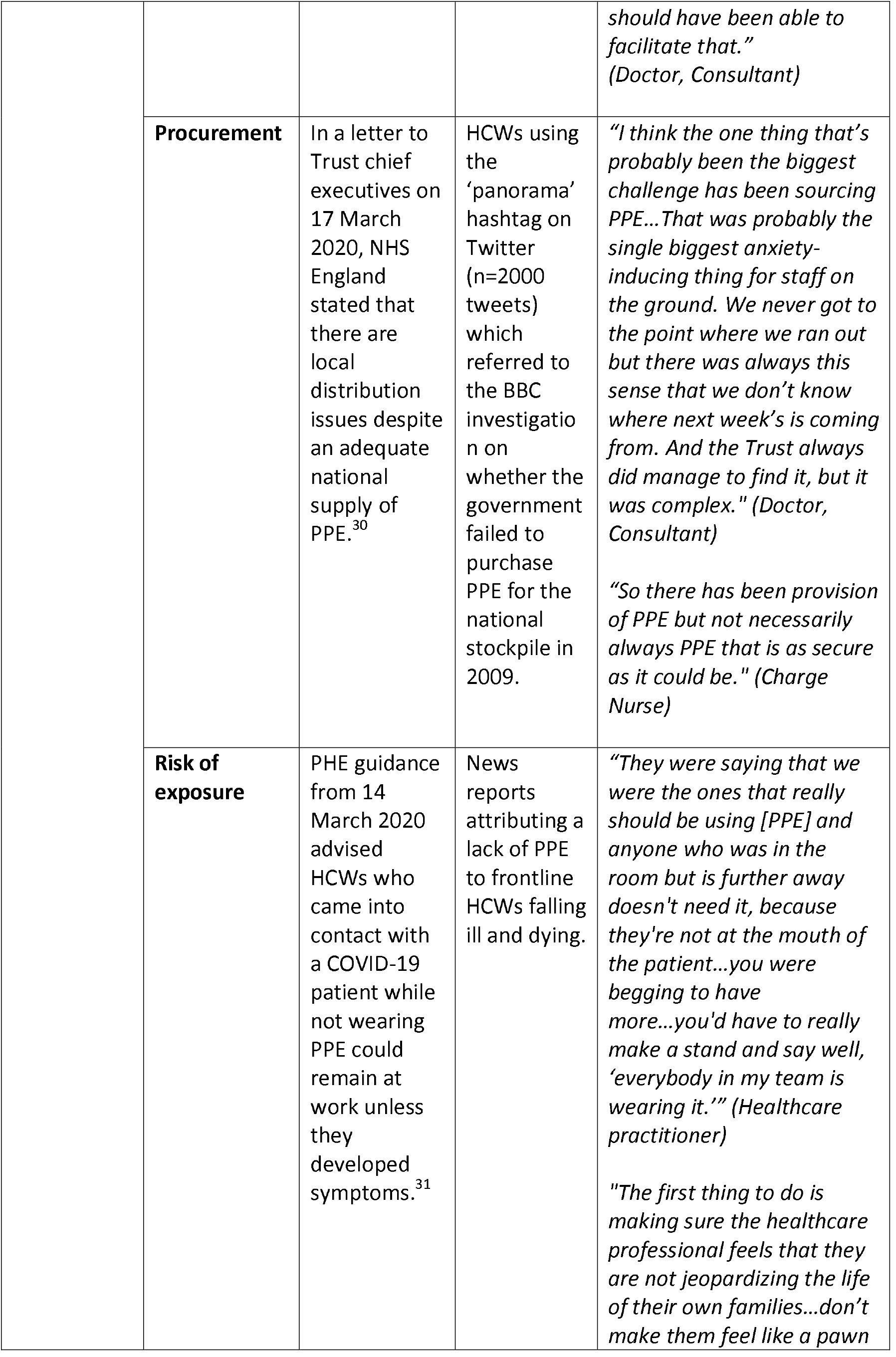
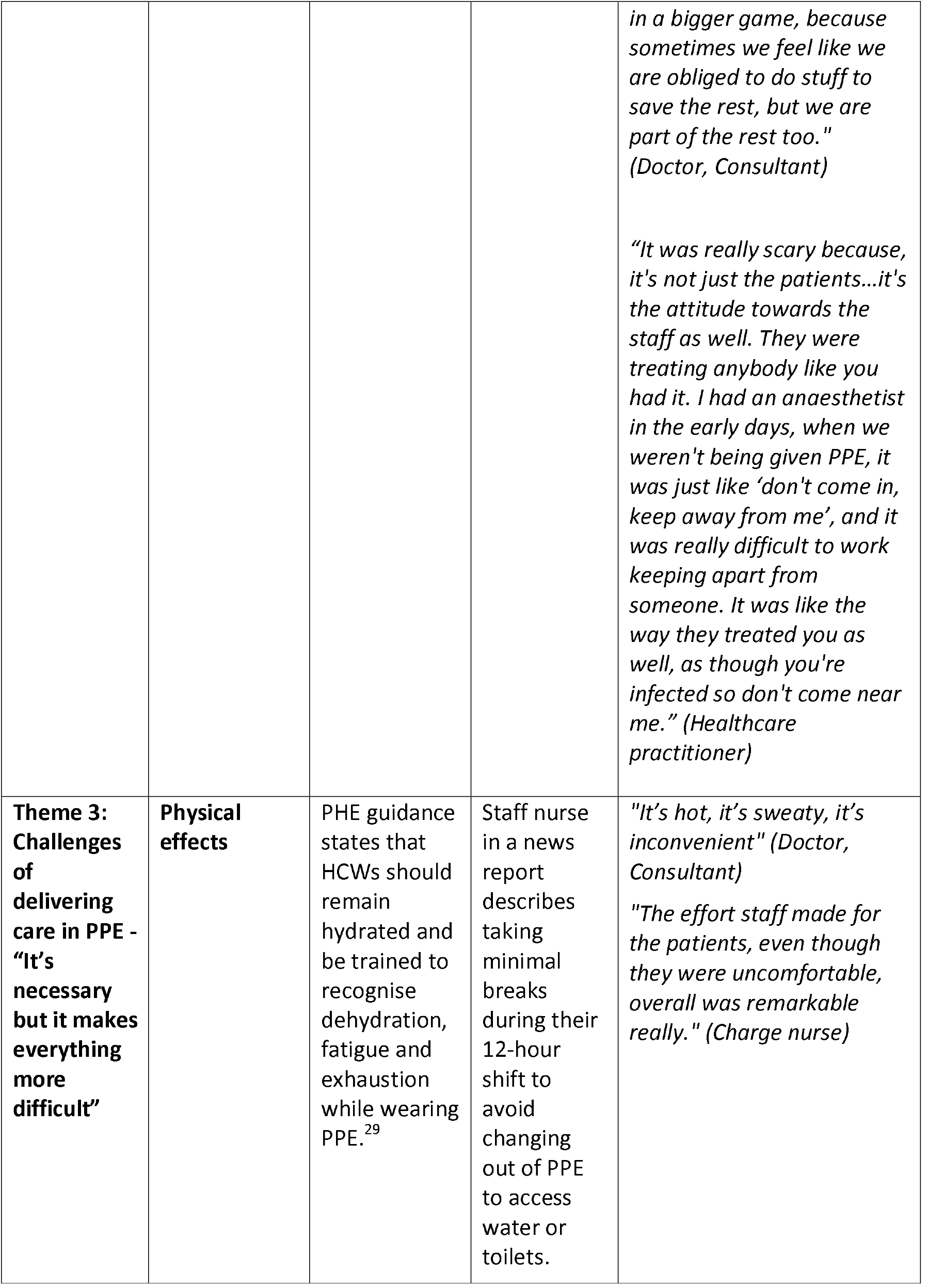
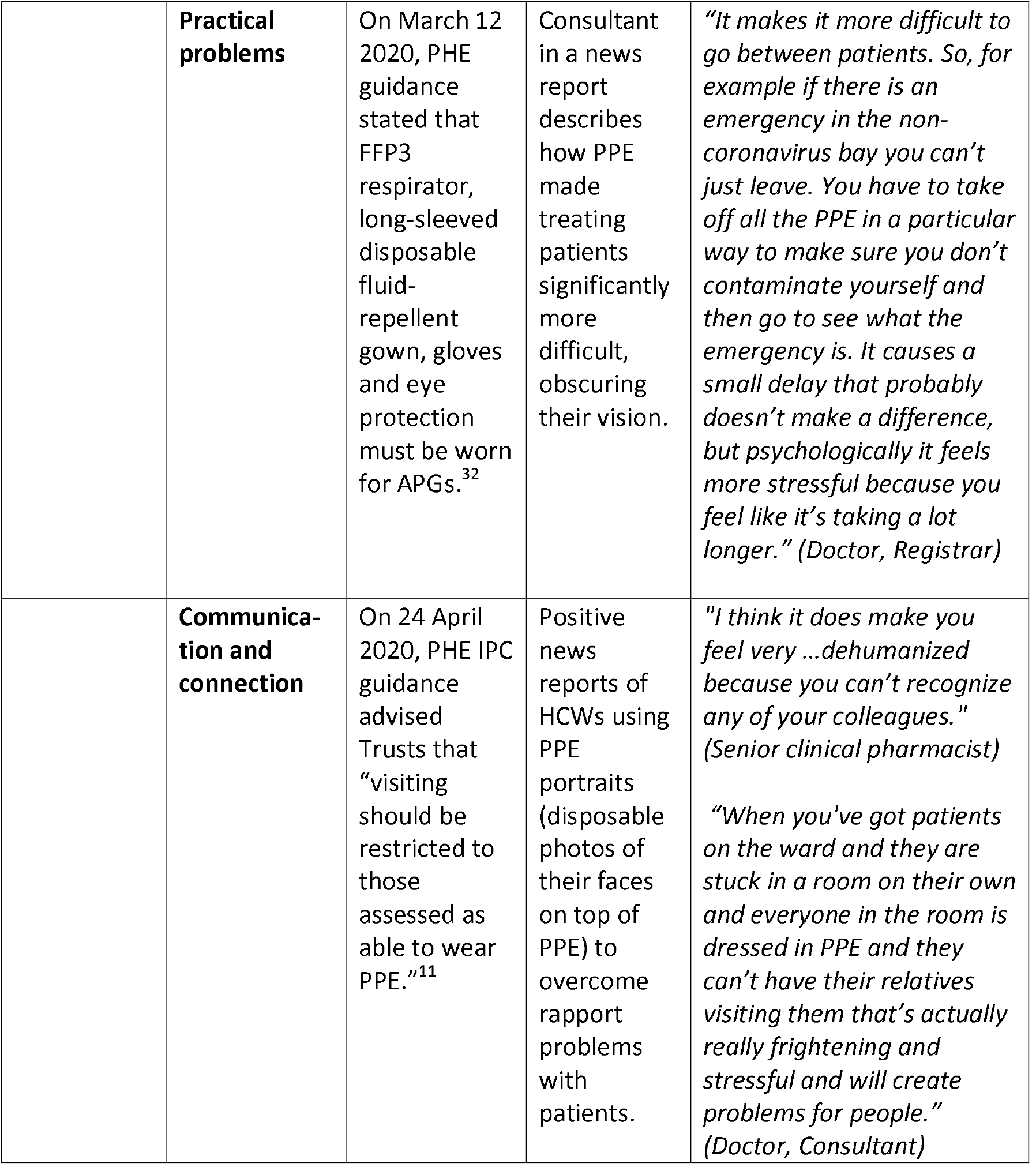
Summary of themes from all streams of data.

### Theme 1: PPE guidance and training - “We weren’t prepared enough”

#### Inconsistent guidance

Towards the start of the outbreak, interviewed HCWs reported limited PPE guidance leading them to care for suspected COVID-19 patients without appropriate PPE. All streams of data analysis found that national PHE and Trust-level PPE guidance changed frequently (see Figure 1), with of daily changes reported in early April 2020. Inconsistent guidance led to confusion, distrust and a lack of confidence in the messaging. HCWs expressed the need for PPE guidance to be more consistent, clearly communicated and provided before HCWs were expected to see patients.

**Figure 1:**
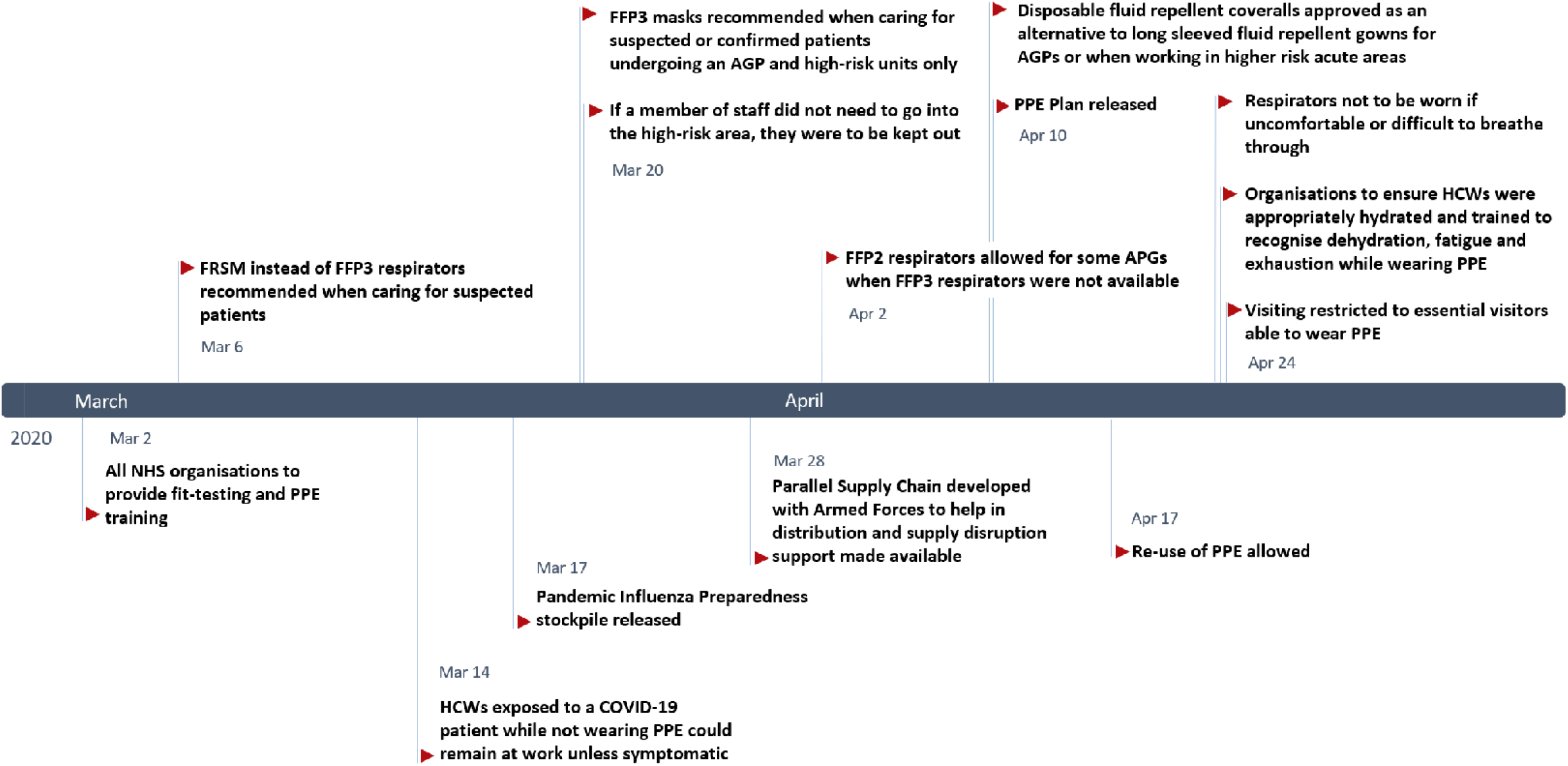
Timeline of changes to national PPE guidance. AGP; Aerosol Generating Procedures, FFP3; Filtering facepiece 3, FRSM; Fluid-Resistant (Type IIR) Surgical Face Mask, HCW; Healthcare worker, High-risk area; ICU, ITU, HDU

On 6 March 2020, PHE recommended that FRSMs were to be used instead of FFP3 respirators when caring for suspected patients.^27^ On 20 March 2020, guidance stated that FFP3 respirators were only needed when managing suspected or confirmed patients, requiring one of their listed “potentially infectious AGPs” and in high risk units such as the ICU, ITU and high-dependency unit (HDU).^33^ On 2 April 2020, guidance changed to advise that if FFP3 respirators were not available, FFP2 respirators could be used instead for some AGPs.^34^ HCWs were concerned that this level of PPE was inadequate. Media analysis showed reports of HCWs being advised to wear single-layer paper surgical masks, instead of FRSMs or FFP3 masks whilst caring for suspected patients. HCWs felt PHE’s list of potentially infectious AGPs^11^ was not comprehensive enough, missing important potential AGPs, such as administering medication via nebulisation and performing chest compressions. HCWs were concerned about the change in PHE guidance on 10 April 2020,^35^ which allowed the use of coveralls with a disposable plastic apron for APGs instead of full-length fluid-repellent gowns. Reports of PPE shortages in interviews and media analyses coincided with the 17 April 2020 PHE guidance which changed to approve the re-use of PPE when there were acute shortages and it was deemed safe to do so.^29^ Having to re-use PPE was distressing, especially when sharing with colleagues. HCWs were concerned that the down-grading and frequent changes to guidance were grounded in supply problems.

As the pandemic progressed, some HCWs felt overwhelmed by increasing amounts of guidance from multiple sources. They felt that having a dedicated team to sort through the information would have increased its clarity. HCWs from community health services found interpreting PPE guidance catered towards hospital-based settings challenging. Senior HCWs were often involved in interpreting national guidance in the context of their local Trust, liaising between staff and management. Some nurses felt as though their voices were not heard in the decision-making processes surrounding PPE guidance and supply on the ward. This was difficult for them as they spent most of their shifts in PPE. HCWs in interviews and the media were concerned about the UK guidance in comparison to other countries, where they felt higher levels of PPE were being provided to HCWs.

#### The training gap

Interviewed HCWs expressed the importance of understanding when, where and how to use PPE. Most interviewed HCWs on ICU, ITU and A&E reported adequate PPE training on how to safely don and doff PPE. However, some HCWs felt there was a “training gap” and expressed the need for earlier, more accessible training available for a wider range of HCWs. A few HCWs reported having had PPE training during past epidemics, but most were unfamiliar with the PPE required for COVID-19 patients. On 2 March 2020, NHS England advised all organisations to provide HCWs with PPE training.^28^ Interviewed HCWs felt PPE training was less accessible to HCWs working outside of high-risk units, such as general wards, surgery, and primary care. Media analysis found training was lacking for HCWs working in the community and in care homes. HCWs took initiative in teaching themselves to safely use PPE when training was not available nor provided early enough. Having training available during both day and night shifts, as well as online materials helped to make PPE training more accessible.

### Theme 2: PPE supply - “If we’re not protected, we can’t protect the public”

#### Shortages (PPE size, level and quality)

HCWs in the media expressed concerns about PPE stockpiles running low from the beginning of March 2020. All streams of data analysis found reports of PPE shortages from across the UK, most notably in care homes, community health facilities and general practice. Visors, full-length fluid-repellent gowns and fluid-repellent facemasks were especially in short supply. One interviewed HCW described PPE being locked in an office with someone monitoring its use. In comparison to intensive care staff, interviewed HCWs from general wards and those from smaller, less prominent hospitals reported greater barriers in access to PPE. Negative sentiment social media posts were mainly related to PPE shortages and a member of parliament (MP) who reported that care homes had adequate PPE. The positive social media were related to deliveries and donations. Informal help and resources advising on appropriate PPE use and how to adapt to limited supplies, was shared on social media.

PHE guidance stated that respirators needed to be the correct size, fit-tested before use, and that HCWs were not to proceed if a “good fit” could not be achieved.^36^ Many HCWs reported failing their respirator fit-test and a lack of alternatives meant that they proceeded caring for COVID-19 patients with these masks or used a lower level of protection. This was especially the case for female HCWs who experienced a lack of small sized masks and scrubs. Media analysis found reports of greater PPE supply problems for BAME HCWs. Powered air purifying respirator (PAPR) hoods (an alternative for HCWs with beards unable to shave for religious reasons) were especially lacking. Concerns were raised that HCWs of lower seniority, including nurses, healthcare assistants and physician associates faced greater barriers accessing PPE. HCWs were also concerned about the quality of PPE. Media analysis found that Trusts, particularly in primary care, received shipments of out-of-date PPE. The policy review found that NHS England stated these shipments of outdated PPE had “passed stringent tests that demonstrate they are safe.”^37^

HCWs reported several adaptions to delivering care in order to preserve PPE, such as the use of open bays with multiple COVID-19 patients, and fewer HCWs seeing patients on ward rounds. Verbal prescriptions were used more frequently to avoid entering the COVID-19 bay and wasting PPE to write a prescription. The policy review found guidance on 20 March 2020 in response to concerns about mask shortages that stated, “if a member of staff does not need to go into the risk area, they should be kept out.”^33^ On 24 April 2020, PHE guidance advised that visiting should be restricted to essential visitors able to wear PPE.^11^ Some HCWs were concerned that PPE supply was a contributing factor limiting families visiting critically ill patients.

#### Procurement

On 17 March 2020, the Department of Health and Social Care (DHSC) announced that there were local PPE distribution problems despite a “currently adequate national supply.”^30^ On 10 April 2020, PHE released their PPE plan which explained that “there is enough PPE to go around, but it’s a precious resource and must be used only where there is a clinical need to do so.”^38^ They emphasised the importance of following national PPE guidance to reduce the significant pressure the supply chain was under. HCWs in interviews and media reported their facilities sourcing PPE at higher costs than usual. Some HCWs resorted to privately purchasing PPE and some Trusts received PPE donations, including 3D printed masks and visors. Extreme examples from the media included HCWs improvising PPE using children’s safety goggles, cooking aprons and bin liners. On social media these concerns were expressed by HCWs using the “panorama” hashtag on Twitter (n=2000 tweets) which referred to the BBC investigation on whether the government failed to purchase PPE for the PIP stockpile in 2009. Even for interviewed HCWs that did not experience PPE shortages, the incremental basis of procurement was concerning for them. HCWs had the perception that, at the local level, facilities should have had prepared larger stockpiles, and that greater international collaboration on global PPE supply chains is needed. Clear communication from Trusts with HCWs about PPE procurement and reassurance that stocks were adequate helped alleviate fears amongst interviewed HCWs.

#### Risk of exposure

Interviewed HCWs feared that a lack of PPE increased their risk of exposure to COVID-19, especially for HCWs that had underlying conditions or were male, BAME, pregnant or been redeployed from retirement. Concerns were compounded by media reports of HCWs in other facilities catching COVID-19 due to insufficient PPE and subsequent exposure to high viral loads. This uncertainty was in the context of a lack of testing for HCWs, causing worries that they were spreading the virus between colleagues, patients and the public. Some HCWs described concerns regarding nosocomial transmission and a change in attitude between colleagues when there was a lack of PPE. A lack of cleaning and changing facilities meant HCWs would wear potentially contaminated clothes home. HCWs expressed concerns about exposing vulnerable household or family members. The policy review found that on 14 March 2020, PHE advised that HCWs who came into contact with COVID-19 patients while not wearing PPE could remain at work unless they developed symptoms.^31^ This policy was subsequently withdrawn on 29 March 2020. HCWs with infectious disease experience, working with adequate provision of PPE and those that had already been ill with COVID-19 reported less fear of exposure. As data collection progressed, HCWs became increasingly used to their new working environments, more familiar with using PPE and less afraid of catching COVID-19.

### Theme 3: The challenges of delivering care in PPE – “It’s necessary but it makes everything more difficult”

#### Physical effects

Interviewed HCWs described PPE to be tiring and uncomfortable to wear, making it more difficult to deliver care. The effects were pronounced for nurses who spent most of their shifts in PPE, and older HCWs with underlying conditions. Tight masks caused facial pain, marks and bruises, rashes, dry skin as well as difficulties breathing, headaches and irritability. HCWs persisted in delivering care despite these effects, often against the PHE advice from 24 April 2020 that respirators “should be discarded and replaced, and not be subject to continued use” when uncomfortable or difficult to breathe through.^11^ For some HCWs, the effects were so severe that they asked to be reassigned to non-COVID wards. Full-length gowns were hot and sweaty, causing overheating and dehydration. Conditions were exacerbated by HCWs fasting during Ramadan and warm weather. HCWs expressed the importance of breaks but often found it difficult to take them, especially on busy wards with shortages of staff and PPE. Wasting PPE on breaks generated feelings of guilt. Drinking less water to avoid having to take breaks made it difficult to follow guidance to remain “appropriately hydrated during prolonged use.”^11^ HCWs expressed the importance of breaks but often found it difficult to take them, especially on busy wards with shortages of staff and PPE.

#### Practical procedures

HCWs found delivering care in PPE to be cumbersome. Donning and doffing PPE contributed to a slower delivery of care, and palpation during physical examinations was less effective with multiple layers of gloves. Goggles fogging up whilst performing procedures, such as intubation and administration of anaesthesia, was frustrating and stressful. Being in PPE restricted HCWs’ movements between patients and wards. Junior HCWs, for example, found that, when in full PPE, they found they were less able to ask for help from seniors outside the COVID-bay not in PPE. HCWs needed to be more prepared than usual when going to see a patient requiring PPE, as they would be unable to leave without doffing and re-donning PPE.

#### Communication and connection

HCWs found it more difficult to build rapport with patients as PPE limited facial expressions, physical touch, and time spent with patients. Being in full PPE could be intimidating, especially for delirious patients. Some HCWs found it difficult to recognise colleagues and often had to shout to be heard through facemasks. Communication problems arose with patients that were elderly and hard of hearing as they relied heavily upon lipreading. HCWs in PPE found alternative forms of communication with colleagues outside of COVID bays, such as portable radios. Some HCWs reported removing their masks when speaking about important topics, such as gaining consent or breaking bad news. HCWs in interviews and media described overcoming rapport problems through use of disposable photos of themselves on their PPE (i.e. disposable photos of their faces attached to gowns).

## DISCUSSION

The findings of this study highlight that HCWs faced multiple challenges when delivering care whilst wearing PPE. Studies from previous epidemics found similar reports of PPE being hot, tiring, time-consuming and restrictive.^39, 40^ In a sample of COVID-19 HCWs with PPE-associated skin problems, Singh et al.^41^ found that 21% took a leave of absence because of this. In addition to the implications for the workforce, they also raised concerns that skin breaches, irritation and increased touching of the face could act as a source of SARS-CoV-2 exposure. HCWs in this study expressed the value of taking breaks to combat the physical effects of PPE but often found it difficult to do so as a result of staff shortages, heavy workloads and guilt over wasting PPE.

PPE reduced HCWs’ ability to develop rapport with patients by masking facial expressions and impairing non-verbal and verbal communication. “PPE portraits” have re-emerged in the COVID-19 pandemic after first being used in the 2014 Ebola outbreak to re-humanise care delivery and have positive anecdotal evidence from HCWs and patients.^42^ Reducing the number of staff on COVID-19 wards to reduce PPE demand raised concerns about increased workloads and quality of care. Objective measures of patient outcomes were beyond the scope of this study but warrant further investigation.

Some participants felt PPE training was not always easily accessible nor implemented early enough. A third of HCWs that responded to a survey by the Royal College of Nurses (RCN) reported on the 8^th^ of May that they had not received PPE training.^5^ Studies on HCWs’ perceptions of working during previous infectious disease outbreaks highlight the importance of PPE training for HCWs to feel confident and prepared to deliver care.^43, 44^ Incorrect use of PPE not only exacerbates shortages but also puts HCWs at higher risk of infection.^45^ HCWs in this study described difficulties accessing training sessions between long shifts and raised concerns that HCWs outside of high-risk settings may experience less training. Previous research has also highlighted that during outbreaks, community HCWs tend to receive less PPE training and face greater difficulties following national guidance often directed towards hospital settings.^46, 47^

There have been widespread reports of UK HCWs experiencing PPE shortages during the COVID-19 pandemic.^48^ Actual and perceived shortages were a major source of anxiety for participants. They advocated for adequate PPE provision to protect their own health and safety. HCWs in China also experienced fears of self-infection and transmission to colleagues, patients and household members due to a lack of PPE.^7^ Changes in guidance to allow the extended use and reuse of PPE also caused distress amongst participants in this study. Evidence on the safety of PPE reuse and extended use is limited, but suggests that it can increase the risk of HCW self-infection and hospital transmission.^45^ This is particularly the case in the absence of clear guidance, protocols and a limited evidence-base on best practice.^49^ Our participants advocated for the need to prioritise adequate provision of PPE for frontline HCWs to protect their own health and safety.

Participants in this study were concerned by the downgrade in guidance from recommending FFP3 respirators to FRSMs,^27^ as well as fluid-resistant full-length gowns to coveralls.^35^ They felt these changes were grounded in supply issues rather than safety measures. Current national guidance may be underestimating the risk of HCWs’ exposure to COVID-19 outside of high-risk settings, potentially resulting in inadequate protection for those HCWs.^49^ Prioritising higher levels of PPE for HCWs in high risk areas is a strategy supported by WHO.^1^ This strategy is thought to have contributed to lower death rates amongst anaesthetists and intensivists.^50^ However, such an approach may be jeopardising the health and safety of HCWs working in lower-risk areas.^51^ PHE guidance recommending FRSMs is lower compared to countries recommending higher level respirator masks (N95, FFP2 or FFP3), such as Australia, USA, China, Italy, Spain, France and Germany.^49^ UK HCWs working on COVID-19 wards following current PHE PPE guidance had nearly three times higher rates of asymptomatic infection compared to HCWs not in COVID-19 areas.^52^ Whilst there are many possible explanations for these findings, an inadequate level of PPE was considered a contributing factor. A key challenge is that research on the transmission of SARS-CoV-2 and the lowest effective level of PPE is ongoing.^53^ Overuse of PPE uses up supplies and may increase risk of transmission through frequent changing, instilling a false sense of safety and potentially reducing the use of other important IPC measures.^54, 55^ However, a recent systematic review and meta-analysis suggests that FFP3 respirators may indeed provide a higher level of protection against infection than FRSMs, even in the absence of AGPs.^56^ HCWs in a study in China experienced no infections with SARS-CoV-2 when provided with appropriate PPE training and supply, including “protective suits, masks, gloves, goggles, face shields, and gowns.”^9^

Participants reported barriers in accessing PPE in general wards, surgery and primary care, as well as in smaller, less prominent hospitals and community clinics. Participants in this study also highlighted that HCWs of lower seniority, female gender and BAME ethnicity may face greater barriers accessing PPE than their colleagues. Female HCWs were concerned about the lack of small sized masks, with some having to deliver care despite failing the fit-test. During the 2015 MERS outbreak in Korea, female HCWs experienced similar difficulties, with oversized coveralls impairing clinical skills and large masks not adequately sealing around their faces, raising concerns about both patient and HCW safety.^57^ Despite only making up 21% of the NHS workforce, BAME HCWs have been overrepresented in the proportion of HCW deaths from COVID-19 in the UK, accounting for 63% of nurses and 95% medical staff deaths.^58^ Official inquiries into the underlying causes of these trends are ongoing.^59^ However, a recent study found that lack of access to PPE was perceived by BAME HCWs in the UK as a major factor contributing to the higher death rates.^60^ Recent studies suggest that in addition to being at greater risk of catching COVID-19, BAME HCWs are more likely to experience inadequate provision and reuse of PPE.^45^ A BMA survey found that only 40% of UK BAME HCWs working in primary care felt they had adequate PPE compared to 70% of white HCWs.^6^ The same survey found that 64% of BAME HCWs felt pressure to work in AGP areas without adequate PPE compared to 33% of white HCWs.^6^

PPE provision for frontline HCWs has become a priority for response efforts across the world. There is an evident need for international collaboration to create sustainable and equitable global PPE supply chains. Many country leaders are calling for equitable and transparent PPE markets to reduce surge in prices and discourage countries bidding against each other.^61^ In the UK, PPE procurement issues existed before the COVID-19 pandemic. The national stockpile was missing critical equipment, such as gowns, which have been short in supply during the pandemic.^62^ A delayed national response, limited domestic PPE manufacturing and exclusion from the EU commission procurement initiatives to secure PPE for its member states left the UK especially vulnerable to shortages.^62^ Knowledge from past epidemics highlights the importance of centralised procurement systems, monitoring PPE use and distributing according to need.^63^

### Implications for practice

HCWs from this study provided insights that can help inform current and future response efforts to the COVID-19 pandemic. Wearing PPE was physically exhausting but HCWs found it difficult to take breaks. Healthcare organisations should provide internal processes to maintain regular breaks for staff working in full PPE, even in contexts of understaffing and PPE shortages as these are key aspects of well-being.^64^ The extra time needed for staff delivering care in full PPE should also be considered when assigning workloads.^54^ Frequently changing guidance generated mistrust towards it. National and Trust-level PPE guidance should be grounded in the most up-to-date evidence base, even in the face of supply problems. Where extended use and re-use of PPE is necessary, comprehensive guidance and training on how to safely do this should be provided. The participants in this study advocated for PPE training to be made accessible to all HCWs from early on to increase preparedness. The findings of this study support that countries should prioritise providing their frontline HCWs with appropriate size, level and quality PPE to avoid perpetuating structural inequalities. Future studies should explore further causes of differential access to PPE in relation to ethnicity, gender and seniority. In the short-term, risk-assessments to identify HCWs at higher risk of COVID-19 and those experiencing barriers accessing appropriate PPE can help ensure targeted procurement is undertaken. This study highlights that Trusts can help alleviate fear amongst their staff by maintaining clear communication about PPE supply and procurement strategies. Adequately prepared stockpiles of PPE for future outbreaks will be critical but international solidarity through equitable and sustainable supply chains will be needed.^65^

### Strengths and limitations

To our knowledge, this is the first qualitative study reporting frontline HCWs’ experiences with PPE during the COVID-19 pandemic in the UK. It offers first-hand experiences from the perspective of HCWs and contributes to the ongoing research on PPE for frontline HCWs during the COVID-19 pandemic.^9^ Interviews were carried out before, during and after the peak of the pandemic, which allowed for experiences to be captured in real-time. However, with the pandemic still evolving, this study will have missed insights since the end of the data collection period. Potential reporting bias should also be considered. Participants may not have been comfortable sharing the full details of their experiences related to PPE during the COVID-19 pandemic, as it became a sensitive and political issue. Participants’ perceived ability to discuss the topic could have been influenced by reports of HCWs being told not to speak to journalists or post on social media about PPE shortages.^66^ The lack of face-to-face contact during interviews limited our ability to pick up on non-verbal cues. Prioritising good rapport with participants was a priority amongst the interview team. This analysis includes data from the initial data collection period, which was limited in its representation of BAME, lower seniority and community HCWs’ experiences. As data collection has progressed, purposive sampling diversified the sample of participants in the wider study.^17^ Triangulating interview findings with media data helped provide insight into the national experience and increase rigour. Although not generalisable to other countries, the UK case offers a lens through which the needs of HCWs’ delivering care on the frontline of the COVID-19 pandemic can be better understood.

## CONCLUSION

This study found that frontline UK HCWs faced multiple challenges delivering care in PPE during the COVID-19 pandemic, including inadequate provision of PPE, inconsistent guidance and a lack of training on its use. Actual and perceived shortages were a large source of distress, creating fears of self-infection and transmission to colleagues, patients and households. Attempting to preserve PPE through reuse and extended use was psychologically and practically challenging. HCWs reported barriers accessing appropriate PPE based on their gender, ethnicity, level of seniority, and healthcare sector. Future research should explore possible reasons for these inequalities. HCWs persisted in caring for their patients despite the negative physical effects, practical problems and communication barriers associated with wearing PPE. HCWs found it difficult to take breaks due to feeling guilty over wasting PPE, busy working environments and lack of protected time for breaks. The findings of this study highlight the importance of protecting HCWs’ health and well-being. In order to feel safe and confident caring for patients, frontline HCWs need to be provided with appropriate size, quality and level of PPE, as well as training in its use. PPE guidance should be consistent, clearly communicated, and reflect the most up-to-date evidence-base for the safest level of PPE. Personal protective needs must be met in order to safeguard the health and well-being of our most valuable healthcare resource: frontline healthcare workers.

## Data Availability

Raw data in the form of interview quotes has been shared and the interview topic guide has been shared.

## Contributors

CVP designed the study, contributed to data collection, supervised data collection, analysis, and reviewed different iterations of the manuscript. KH, LM, ND and SLJ contributed to the interviews. KH, LM and SLJ conducted the policy review. SM and SV contributed to social media monitoring, data collection and analyses. KH conducted data analysis of all data and wrote the first draft of the manuscript. ND supervised data analysis and reviewed different iterations of the manuscript. All authors edited the manuscript and approved the final version.

## Transparency statement

The lead author and guarantor (KH) affirms that this manuscript is an honest, accurate, and transparent account of the study being reported; that no important aspects of the study have been omitted; and that any discrepancies from the study as planned (and, if relevant, registered) have been explained.

## Patient and Public Involvement

The study protocol and study materials were reviewed by the team’s internal patient and public involvement panel. The panel’s feedback was used to make changes in the research questions and study materials.

## Ethical approval

This study was approved by the Health Research Authority (HRA) and Health and Care Research Wales (HCRW) and the R&D offices of the hospitals where the study took place. IRAS project ID: 282069

## Trial registration details

Not applicable as this was not a clinical trial.

## Dissemination statement

We plan to disseminate the results to study participants.

## Acknowledgments

This study was conducted by the Rapid Research Evaluation and Appraisal Lab (RREAL) at UCL. We would like to thank the support provided by Anna Dowrick, Anna Badley, Caroline Buck, Norha Vera, Nina Regenold, Kirsi Sumray, Georgina Singleton, Aron Syversen, Elysse Bautista Gonzalez, Lucy Mitchinson, Harrison Fillmore and Anna Badley.

## Appendices

Appendix 1: Inclusion criteria and search terms

|  |  |  |
| --- | --- | --- |
| **Inclusion criteria** | **Newspaper data** | 1) Published between 15 March 2020 and 5 June 2020;<br>2) Aimed at healthcare delivery;<br>3) Related to the COVID-19 pandemic;<br>4) Related to personal protective equipment. |
|  | **Social media data** | 1) Published between 1 December 2019 and 31 May 2020.;<br>2) Aimed at healthcare delivery;<br>3) Related to the COVID-19 pandemic;<br>4) Related to personal protective equipment. |
|  | **Policy data** | 1) Published between 1 December 2019 and 5 June 2020.;<br>2) Aimed at healthcare delivery;<br>3) Related to the COVID-19 pandemic;<br>4) Related to personal protective equipment. |
| **Search terms** | **Media and policy data** | COVID-19 OR coronavirus OR corona |
|  | **Social media data** | ((bio:"healthcare professional" OR bio:"healthcare worker" OR bio:"doctor" OR bio:"NHS" OR bio:"nurse" OR bio:"physio*" OR bio:"Paramedic" OR bio:"Ambulance work*" OR bio:"Ambulance driver*") AND ("coronavirus" OR "#coronavirus" OR "corona" OR "COVID-19" OR "COVID 19" OR "COVID19" OR "#COVID19" OR "COVID_19" OR "COVID" OR "severe acute respiratory syndrome coronavirus 2" OR "severe acute respiratory syndrome coronavirus 2" OR "2019-nCoV" OR "SARS-CoV-2" OR "2019nCoV" OR "physio*" OR "PPE") OR ("i am" OR "as a" OR "source: I" OR "I'm a") near/5 ("doctor" OR "nurse" OR "doctors" OR "nurses" OR "Paramedic" OR "Ambulance worker" OR "Ambulance driver") AND ("coronavirus" OR "#coronavirus" OR "corona" OR "COVID-19" OR "COVID 19" OR "COVID19" OR "#COVID19" OR "COVID_19" OR "severe acute respiratory syndrome coronavirus 2" OR "severe acute respiratory syndrome coronavirus 2" OR "2019-nCoV" OR "SARS-CoV-2" OR "2019nCoV" OR "physio*" OR "PPE") NOT ("I am not" OR "I'm not")) |

